# Multitrait Genome-Wide Analysis in the UK Biobank Reveals Novel and Distinct Variants Influencing Cardiovascular Traits in Africans and Europeans

**DOI:** 10.1101/2022.02.27.22268990

**Authors:** Musalula Sinkala, Samar Elsheikh, Mamana Mbiyavanga, Joshua Cullinan, Nicola Mulder

## Abstract

In exploring the trans-ancestral genetic nuances of cardiovascular traits, we conducted a multi-trait genome-wide association study focusing on African (AFR) and European (EUR) populations in the UK Biobank. Here, we identify 50 genomic risk loci in the AFR population, among which 43 are novel discoveries associated with four cardiovascular traits. Similarly, we identify 829 loci in the EUR population, with 47 being novel. Also, at these loci, we identify 52 SNPs (45 novel) in the AFR population and 1,856 SNPs (957 novel) in the EUR population, among which 83 are shared, highlighting both the shared and rich diversity of the genetic underpinnings of cardiovascular disease across populations. Furthermore, functional mapping of these SNPs reveals distinct distribution patterns, with the EUR population showing a higher proportion in intronic and untranslated regions. Further, our study unravels population-specific genetic associations, identifying 3,011 genes exclusive to the EUR group and 36 distinct to the AFR group. Additionally, gene enrichment analyses show unique enriched pathways for each population, highlighting the potential influence of genetic ancestry on cardiovascular trait mechanisms and manifestation. Collectively, our results underscore the importance of population-specific approaches in studying the genetic underpinnings of cardiovascular health and further indicate potential avenues for personalised medicine and targeted interventions.

## Introduction

Cardiovascular traits encompass a complex spectrum of phenotypes dictated by a confluence of genetic and environmental determinants^1,2^. Intriguingly, the distribution of these cardiovascular indices exhibits striking disparities across various ancestral backgrounds^3,4^. In recent years, Genome-wide association studies (GWAS) have illuminated numerous genomic loci associated with these cardiovascular traits ^5–7^, highlighting the intricate genetic fabric that underpins cardiovascular health. Traits such as systolic blood pressure (SBP), diastolic blood pressure (DBP), and heart rate share loci with pathologies like stroke and myocardial infarction, and these associations often serve as harbingers of early mortality and general health^8–11^. For instance, seminal investigations identified 535 novel variants linked with blood pressure traits in over a million European individuals^7^, while another study unearthed hundreds of systolic and diastolic BP-associated variants from the UK Biobank cohort^12^. Crucially, most of these findings stem from populations of European descent^13^.

Despite bearing the highest hypertension burden, the African population remains comparatively under-explored in GWAS^13,14^. Emerging large-scale GWAS and meta-analyses are beginning to shed light on the genetic underpinnings of this complex condition within this and other populations^15–18^. However, few extensive studies have concurrently examined the genetic and social health determinants, despite compelling evidence substantiating their contribution to hypertension^12^.

Drawing on evidence from a host of GWAS investigations into other traits^19–25^, we posit that genetic variants influencing cardiovascular traits may manifest divergently among European and African individuals. Elucidating such variances may shed light on the differential genetic underpinnings and pathophysiological discrepancies between these demographic groups observed in cardiovascular traits and disease risk^14,15,26^. However, despite the significant advances in multi-trait GWAS, there is a distinct paucity of research efforts focused on identifying and contrasting genomic loci associated with cardiovascular traits in African and European populations (note, here we use these terms to refer to populations of European and African descent, respectively, rather than those necessarily living in these geographical regions) using multiple cardiovascular measurement parameters.

This study explores the heterogeneities in cardiovascular traits, encompassing SBP, DBP, pulse rate, and ‘maximum heart rate during fitness Test’ (MHR), between African and European individuals represented in the UK Biobank^27^. We performed multi-trait genome-wide association analyses (MTAG) to discern sets of variants influencing these cardiovascular traits (SBP, DBP, pulse rate, and MHR) within each population group. Finally, leveraging integrative enrichment analyses, we sought to identify the associated genes, pathways, and phenotypes. Collectively, our findings reveal distinct genetic determinants influencing cardiovascular traits in African and European populations, thereby advancing our understanding of the genetic intricacies of cardiovascular health across diverse populations.

## Methods

### Data Collection and Demographic Selection

We analysed datasets derived from the UK Biobank^27^, including genotyping array data, SBP, DBP, pulse rate, MHR, and additional anthropometric measurements. Our investigations were constrained to a subset of these data, comprising 460,096 individuals of European descent (classified as White, British, Irish, or “any other White background”) and 6,551 individuals of recent African ancestry (classified as Black). The specifics of UK Biobank participant selection, recruitment methodologies, and sample collection and analysis protocols have been comprehensively documented in prior publications^27,28^.

### Comparison of cardiovascular traits in Europeans and Africans

We compared the mean values for SBP, DBP, pulse rate, and MHR between the European (n=460,096) and African (n=6,551) participants. We applied a student’s t-test for these comparisons, assuming unequal variances. To further probe the relationships between these cardiovascular measures and other anthropometric traits, we calculated Pearson’s correlation coefficients, providing a measure of the linear dependencies between these variables. Finally, to discern the impact of participant BMI on DBP, we visualized the trend via plotting DBP over each 10th percentile bin, with error bars included to denote uncertainty.

### Genome-wide identification of genetic loci associated with cardiovascular traits

We collected GWAS summary statistics of cardiovascular traits, namely SBP, DBP, pulse rate and MHR, of 396,670 Europeans and 6,551 Africans profiled by the UK Biobank. The approach towards participant genotyping within the UK Biobank has been thoroughly detailed in previous reports^27,29^. Further information regarding the quality control measures employed in the genotyping analyses can be accessed at https://pan.ukbb.broadinstitute.org/docs/qc. GWAS summary statistics, as calculated by the UK Biobank project for each cardiovascular trait, were used in our investigation.

The GWAS protocol applied to the cardiovascular and ancestry groups, and the specifics of the analytical techniques are extensively documented in earlier publications^30^. The procedure, in brief, implemented the Scalable and Accurate Implementation of the Generalized Mixed Model approach^31^. A linear or mixed logistic model was adopted, incorporating a kinship matrix as a random effect and certain covariates as fixed factors. The covariates incorporated in the model include the participant’s age and sex, the interaction term of age and sex, squared values of age, the interaction of squared age and sex, and the initial ten principal components derived from the genotype datasets.

We processed these summary statistics through a two-stage approach. In the first stage, we conducted separate single GWA for African and European populations, examining associations with SBP, DBP, pulse rate and MHR.

In the second stage, we applied multi-trait analysis of GWAS (MTAG) across all four traits to boost the discovery power for SNPs and investigate the genetic underpinnings of crucial cardiovascular traits. Utilising summary statistics from the UK Biobank, we analysed AFR and EUR populations separately, thereby capturing population-specific genetic variation. The MTAG analyses were implemented through the METAL software tool^32^, adhering to default settings to ensure the consistency and replicability of the findings. The creation of Manhattan plots was facilitated using MATLAB, utilising the software delineated in prior literature^33^.

### Fine-Mapping and Functional Annotation of Genetic Associations

Intending to identify independent SNP associations and genomic risk loci, along with mapping the consequential genes, we employed Functional Mapping and Annotation (FUMA)^34^, an integrative platform for functional annotation, for our subsequent analyses. This was executed utilising the MTAG summary statistics, distinctly for the AFR and EUR populations.

We applied FUMA to facilitate a comprehensive interpretation of the GWA results. The analysis pipeline started with the independent SNP identification based on the single trait and multi-trait GWA results. Then, applying the default settings, SNPs with a p-value < 5 × 10^−8^ and an LD r2 > 0.1 in the AFR or EUR population separately, SNP level filtering to exclude SNPs with minor allele frequency < 0.01 and were acknowledged as independent. Post-identification, these independent SNPs were aligned into genomic risk loci.

Following the SNP-based analysis, gene mapping was conducted, prioritizing the genes that the identified SNPs could potentially influence. The purpose was to comprehend how genetic variations at these loci might induce changes in cardiovascular trait regulation. Further, we employed FUMA’s functional annotation capabilities to assess the functional impact of these identified SNPs. This provided a comprehensive annotation of the functional consequences of the variants, aiding in understanding their potential biological relevance.

In summary, the exhaustive analyses in FUMA, employing the MTAG summary statistics distinctly for AFR and EUR populations, encompassed the identification of lead and independent SNPs, delineating genomic risk loci, assessing the functional impact of SNPs and mapping related genes.

### Categorisation of novel and known variants leveraging LDlink and FUMA

In our study, we identified novel and reported (known) variants from LDLink ^35^, an online suite of applications that provides easy access to population-specific LD information. This was accomplished by separately utilising the list of lead SNPs from the Multitrait GWAS with cardiovascular traits in AFR and EUR populations.

The independently significant SNPs were first obtained from FUMA outputs for the AFR and EUR analysis, and then we automated retrieving data from LDlink’s RESTful API using a cURL command for each SNP. In this query, each SNP was tested against the GWAS Catalog^36^ using the LDtrait software^37^, which provides information on known trait associations. Then, in the context of the AFR and EUR populations, we applied the respective AFR and EUR reference panels in LDlink to identify comparable variants. The approach was optimized by employing a window size of 500 kilobase pairs and implementing an r2 threshold of 0.1 for LD calculations. We deemed SNPs novel if there were no previously reported significant variants in the GWAS Catalog or previous studies reported in LDlink that fall within ±1Mb from the lead variant.

The significant lead SNPs we identified associated with cardiovascular traits were classified into three categories: ‘Novel’, ‘Has Trait’, and ‘Cardiovascular Risk Associated’. The ‘Novel’ category comprises SNPs not previously identified in the GWAS Catalog or the literature. The ‘Has Trait’ category included SNPs associated with specific traits. Finally, the ‘Cardiovascular Risk Associated’ category, designed per the secondary analysis, included SNPs directly associated with cardiovascular or systemic traits like body weight and BMI.

### Cross-Population Replication of Cardiovascular Trait-Associated Variants

To deepen our understanding of population-specific genetic underpinnings, we sought to replicate the significant findings of cardiovascular traits from EUR within AFR populations and vice versa.

Our replication methodology centred on the examination of linked variants. For lead variants that displayed a significant association with cardiovascular traits, we identified all associated variants within an LD block (r^2^ > 0.1) extending ±500 KB from the lead variant. We then assessed the association of these linked variants within the EUR population with the same trait within the AFR population and vice versa. This was achieved by extracting the corresponding GWAS p-values from the AFR dataset.

To account for multiple testing and control the false discovery rate, we adjusted these p-values using the Benjamini and Hochberg correction procedure^38^. Variants within each LD block that showed a statistically significant association (adjusted p-value < 0.05) and shared the same direction of effect (either positive or negative) as the lead variants were considered indicative of local replication.

### Identification and Functional Annotation of Cardiovascular Trait-Associated Genetic Variants

We individually analysed the AFR and EUR cohorts, extracting genetic variants related to cardiovascular traits from the MTAG summary statistics. Following this extraction, we applied established variant-to-gene mapping techniques in FUMA to elucidate the genes connected to these variants in both populations. Our next step was to assess the enrichment of the mapped genes, employing the Enrichr^39^ platform. We explored a variety of databases - ClinVar^40^, DisGeNET^41^, WikiPathway^42^, and UK Biobank Gene set term defined using GWAS p-values, computed by Maya’an Lab^39^, for an array of UKB phenotypes^39^, - for this purpose. This comprehensive strategy enabled us to identify enriched annotation terms, further refining our understanding of the biological implications of our findings.

### Statistics

We employed a suite of computational tools, including R, Python, MATLAB 2021a, and Bash, to perform our statistical analyses. Our established significance threshold was a two-sided p-value of less than 0.05 for individual comparisons. In addition, to account for the potential inflation of the type I error rates inherent in multiple comparisons, we implemented the Benjamini & Hochberg procedure^38^ to control the false discovery rate, thus ensuring the robustness of our findings.

## Results

### Distinct cardiovascular profiles characterise African and European populations

In our exploration of the UK Biobank datasets, encompassing cardiovascular metrics from 460,096 Europeans and 6,551 Africans, we noted a significant distinction in the mean diastolic blood pressure (DBP). Africans exhibited a markedly higher mean DBP of 84.7 mmHg, compared to 82.0 mmHg in Europeans (Welch test: t=13.9, p=1.2 × 10^−42^; Figure 1a). In contrast, there was no significant difference observed in the mean systolic blood pressure (SBP) between Africans (mean=139.8 mmHg) and Europeans (139.4 mmHg), p=0.187; Figure 1b).

**Figure 1:**
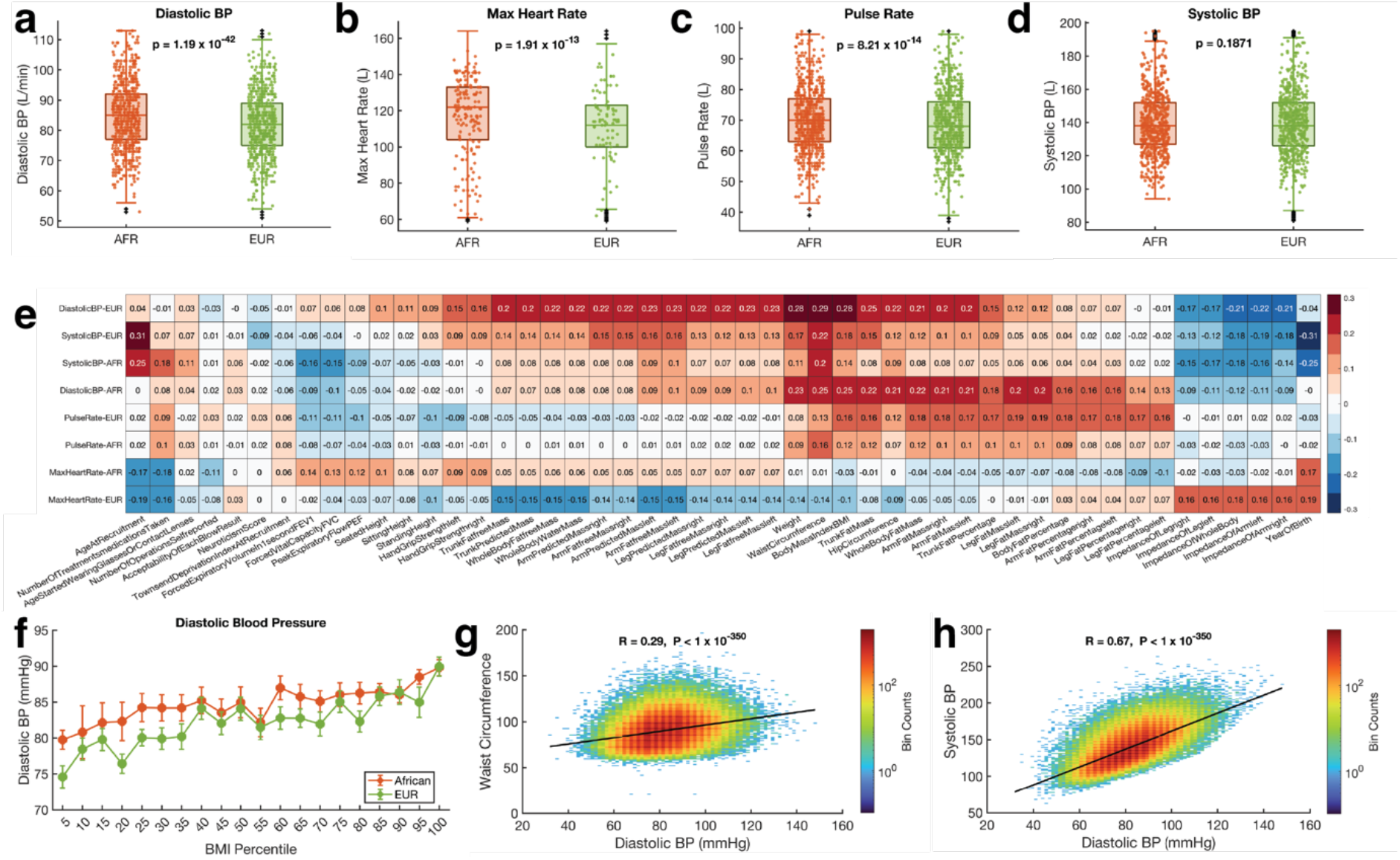
Cardiovascular Parameters and Anthropometric Correlations in Africans and Europeans: Comparison of the cardiovascular function parameter in Africans and Europeans for **(a)** diastolic blood pressure, **(b)** systolic blood pressure, **(c)** pulse rate, and **(d)** maximum heart rate during fitness test.

The boxplots indicate the distribution of each measurement among Africans and Europeans. The p-values shown for each comparison were calculated from Welch’s t-test. On each box, the central mark indicates the median, and the left and right edges of the box indicate the 25th and 75th percentiles, respectively. The whiskers extend to the most extreme data points not considered outliers, and the outliers are plotted individually using the ‘+’ symbol. To make the visualisation clearer, the filled circle mark showing the distribution only include 1000 randomly sampled point from the entire samples size of each group. **(e)** Correlation between SBP, DBP, pulse rate, and MHR with other anthropometric measurements in Africans and Europeans. **(f)** Error bars show the DBP variation across BMI percentiles among Africans and Europeans. The middle point indicates the mean DBP, and the error bars indicate the standard error of the mean at the particular BMI percentile. Binned scatter plot showing Pearson’s linear correlation between **(g)** DBP and waist circumference and **(h)** DBP and SBP. The data points are spaced into rectangular bins, and each point is coloured based on logarithm bin size, with redder colours indicating a higher number of plots. The colour bar shows the colour scale.

Furthermore, our investigation revealed significant differences in pulse rate and MHR across the two populations. Africans consistently exhibited higher values (mean pulse rate=70.3 bps, mean MHR=116.0 bps) than their European counterparts (pulse rate=68.9 bps, MHR=110.3 bps). The statistical analysis highlighted these differences with p-values of 8.2 × 10^−14^ for pulse rate (Figure 1c) and 1.9 × 10^−13^ for MHR (Figure 1d), further highlighting the distinctive cardiovascular profiles between these two populations.

Existing literature indicates that SBP, DBP, and pulse rate exhibit considerable variations in relation to age and other anthropometric measures of individuals^43–46^. Therefore, we conducted a rigorous assessment of the linear relationship, using Pearson’s correlation, between the cardiovascular traits in conjunction with a series of anthropometric measurements (Figure 1d, Figure 1e and Figure 1f). Here, our analysis revealed a positive linear correlation between the SBP and DBP of the participants (Figure 1g and 1h). The mean comparison test statistics and descriptive statistics of the EUR and AFR individuals are summarised in Supplementary Data 1.

### Multi-trait genetic analysis reveals robust variations in cardiovascular traits between African and European populations

We began with an initial analysis utilising GWAS summary data from the UK Biobank. These encompassed variants tied to four cardiovascular traits - SBP, DBP, pulse rate, and MHR, from two distinct ethnic cohorts: Africans (AFR; n=6,551) and Europeans (EUR; n=396,670). For the AFR population, we found 72 different candidate SNPs that were significantly associated (using a genome-wide significance threshold of p<5 × 10^−8^) with Pulse Rate, which, after accounting for linkage disequilibrium (LD) using analysis in FUMA returned only two independent SNP associations, rs9388010 (p=6.4 × 10^−12^) and rs9375066 (p=1.4 × 10^−10^) (see Supplementary Table 1). No significant associations were observed for Systolic BP, Diastolic BP, or MHR, most likely due to the relatively small sample size.

In contrast, for the EUR population, our analysis identified a total of 3,365 independent SNPs associated with Pulse Rate, 2,763 independent SNPs associated with Systolic BP, 2,988 independent SNPs associated with Diastolic BP, and 36 independent SNPs associated with Max Heart Rate (MHR) (Supplementary Table 1).

Next, we took advantage of the correlation between the cardiovascular traits to boost the power for the discovery of SNPs by performing a multi-trait analysis (MTAG) as the second stage of our GWA analysis combining all four traits (SBP, DBP, Pulse Rate, and MHR). The MTAG served to integrate findings across all four traits.

Our study identified 829 genomic risk loci for the EUR population, encompassing 1,856 lead SNPs (SNP in the locus that shows the most significant association with the trait of interest) and 5,416 individually significant (MTAG p < 5 × 10^−5^) SNPs (Figure 2a). These findings were derived from a pool of 138,032 candidate SNPs, with 134,477 of these SNPs supported by prior GWAS findings, strengthening their link to various traits (see Supplementary Table 2, Supplementary Data 2).

**Figure 2:**
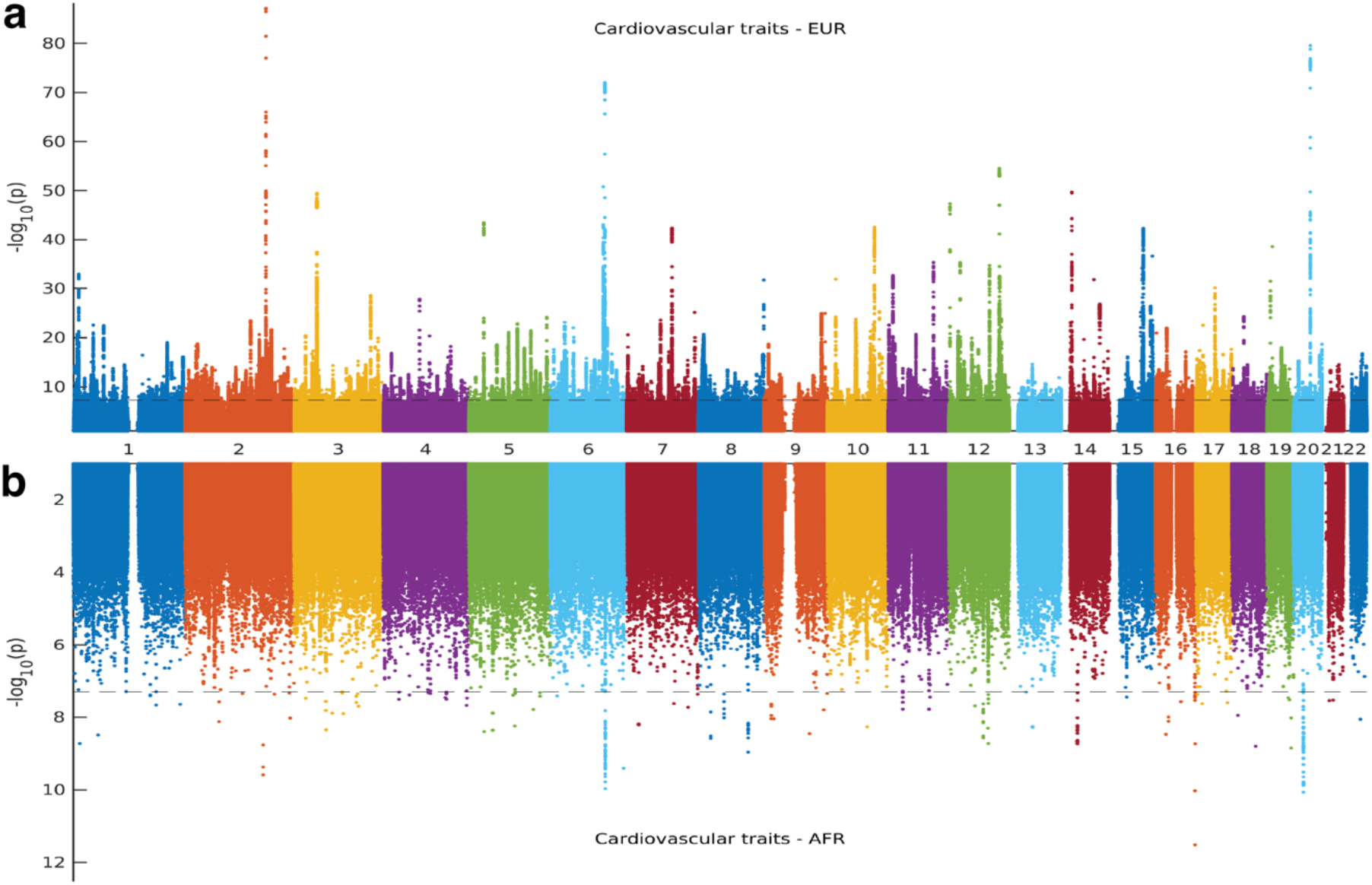
Manhattan Plots of Cardiovascular Trait-Associated Variants in European and African Populations: plots representing MTAG analysis variants associated with cardiovascular traits (DBP, SBP, Pulse Rate, and MHR) in European and African populations for each chromosome.

Additionally, we mapped these candidate SNPs to 3,033 genes that may mediate the observed associations (Figure 2a).

In contrast, the analysis in the AFR population revealed 50 genomic risk loci, comprising 52 lead SNPs and 54 individually significant SNPs (Figure 2a). These findings emerged from 772 candidate SNPs, of which 753 were tagged by previously reported GWAS findings, confirming the role of these loci in various traits within this population (see Supplementary Table 2 and Supplementary Data 2). We linked these candidate SNPs to 46 genes that may contribute to the observed associations.

A total of 403,221 samples for GWA from the EUR and AFR populations were analysed in this study. The EUR population had a significantly larger sample size (396,670) than the AFR population (6,551). Thus, the AFR sample size was roughly 60 times smaller than the EUR sample size. The data were evaluated for four traits using MTAG across both populations, which amounted to 26,204 observations in AFR and 1,586,680 in the EUR population. The larger sample size in the EUR population led to the detection of more SNPs^47,48^, while the smaller AFR sample size most likely accounts for the fewer SNP-trait associations detected.

### Distinct population-specific variants underlie cardiovascular trait differences between Africans and Europeans

Taking into consideration the unique LD structures of AFR and EUR populations, we embarked on a comparative analysis of the genomic risk loci and SNPs associated with cardiovascular traits in these distinct demographic groups (see Methods section).

Among the 50 loci identified in the AFR population, we successfully replicated all of them in the EUR population. Conversely, out of the 829 loci identified in the EUR population, we replicated 154 loci in the AFR population, revealing a shared genetic risk among individuals of AFR and EUR ancestry (see Supplementary Data 2). These findings highlight the existence of shared genetic architecture contributing to disease susceptibility in diverse populations.

From a set of 1,856 lead SNPs that we identified within the EUR population, we replicated 60 in the AFR population. Notable replicated SNPs include rs143362094 (EUR MTAG p=2.0 × 10^−08^; AFR MTAG p=2.8 ×10^−10^), rs57182997 (EUR p=4.4 × 10^−13^; AFR p=1.3 × 10^−09^), and rs9482195 (EUR p=7.9 ×10^−09^; AFR p=7.1 × 10^−09^) (Supplementary Data 2). Conversely, from 52 individually significant SNPs pinpointed in the AFR population, 29 were replicated within the EUR population, including rs9388008 (AFR MTAG p=1.1 × 10^−10^; EUR MTAG p=1.8 × 10^−72^), rs77199928 (AFR p=1.9 × 10^−08^; EUR p=4.0 × 10^−16^), rs10069648 (AFR p=3.8 × 10^−08^; EUR p=1.6 × 10^−13^) (see Table 1, and further details in Supplementary Data 2).

Our analysis revealed many SNPs associated exclusively with either the EUR or AFR populations. Specifically, we found 1,795 SNPs, including rs4811602 (MTAG EUR p=2.8 × 10^−80^) and rs10774625 (p=2.9 ×10^−55^), associated only with the EUR population and 29 SNPs, including rs200399297 (MTAG AFR p=1.6 × 10^−09^), rs148281566 (p=1.9 × 10^−09^) and rs17055585 (p=2.6 × 10^−09^), specific to the AFR population. The AFR-specific and EUR-specific SNPs are detailed in Supplementary Data 2.

**Table 1:** Top variants associated with cardiovascular traits in AFR and EUR.

| Ethnicity | uniqueID | LeadSNP | LeadSNP_P | LinkedSNP | Rsquare | Zscore_EUR | Zscore_AFR | LinkedSNP_P |
| --- | --- | --- | --- | --- | --- | --- | --- | --- |
| AFR | 6:122272775:C:T | rs143362094 | $2.0 \times 10^{-8}$ | rs2091624 | 0.1224 | -18.0 | -6.3 | $2.8 \times 10^{-10}$ |
| AFR | 6:122061840:C:CA | rs57182997 | $4.4 \times 10^{-13}$ | rs9388004 | 0.1841 | 14.1 | 6.1 | $1.3 \times 10^{-9}$ |
| AFR | 6:122249264:G:T | rs9482195 | $7.9 \times 10^{-9}$ | rs1919865 | 0.1036 | 16.1 | 5.8 | $7.1 \times 10^{-9}$ |
| AFR | 4:3089564:C:T | rs61348208 | $5.0 \times 10^{-10}$ | rs910568 | 0.2693 | 3.1 | 5.3 | $1.1 \times 10^{-7}$ |
| AFR | 5:68414152:A:G | rs351433 | $4.1 \times 10^{-10}$ | rs2076823 | 0.1065 | 2.5 | 5.1 | $4.1 \times 10^{-7}$ |
| EUR | 6:122138479:A:C | rs9388008 | $1.1 \times 10^{-10}$ | rs9401451 | 0.2555 | -18.0 | -4.6 | $1.8 \times 10^{-72}$ |
| EUR | 5:157878049:A:C | rs10069648 | $3.8 \times 10^{-8}$ | rs2419922 | 0.1739 | 7.4 | 2.0 | $1.6 \times 10^{-13}$ |
| EUR | 3:71045995:A:T | rs9853237 | $4.5 \times 10^{-9}$ | rs66590721 | 0.1219 | 5.8 | 2.8 | $7.6 \times 10^{-09}$ |
| EUR | 12:87006555:A:G | rs77304792 | $2.4 \times 10^{-8}$ | rs1922749 | 0.2068 | 4.4 | 1.9 | $1.0 \times 10^{-6}$ |
| EUR | 1:184415295:A:G | rs549702399 | $2.2 \times 10^{-8}$ | rs12726204 | 0.2035 | 4.4 | 3.5 | $1.2 \times 10^{-5}$ |

Moreover, our findings identified 29 AFR-specific SNPs and 1,795 EUR-specific SNPs that were not successfully replicated in the opposite population. Finally, we observed 83 common variants between AFR and EUR, accounting for the SNPs in LD with significant SNPs in each group (see Supplementary Figure 1).

Next, we conducted a comparative analysis of the beta estimates of candidate single SNPs to evaluate their effect sizes and provide further insights into the distinctiveness of variant effects. Notably, SNPs that were significantly associated with cardiovascular traits in the AFR cohort exhibited a larger effect size within this group compared to the EUR population and vice versa (Figure 3a, Figure 3b).

**Figure 3:**
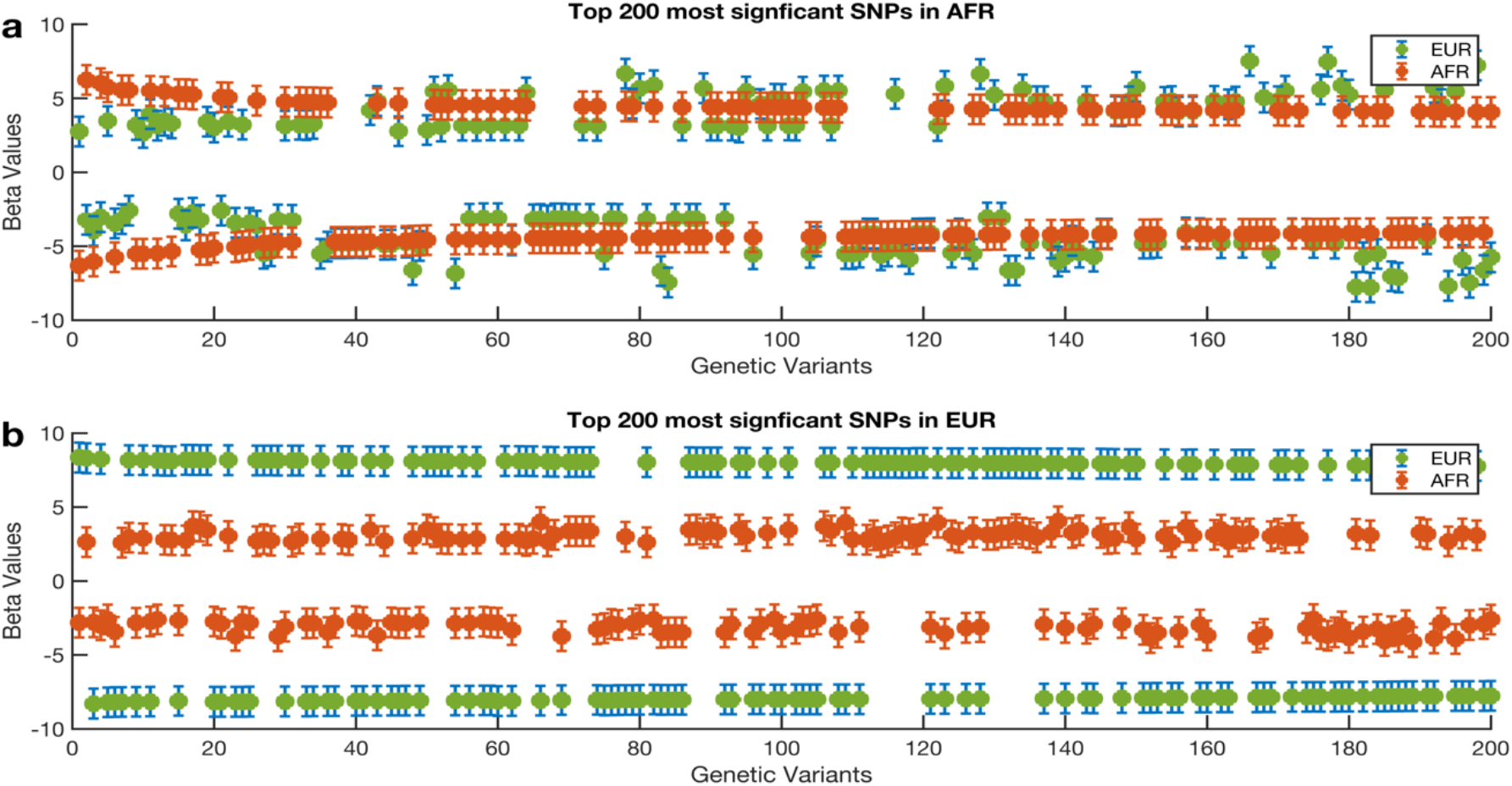
Comparison of Beta Estimates for Top 200 Cardiovascular Trait-Associated Variants: The 200 most significant variants associated with cardiovascular traits in (a) African and (b) European populations. The y-axis illustrates beta estimates and associated errors derived from MTAG analyses, estimated from the z-score output of METAL. The central point of each error bar represents the beta estimate, while the whiskers demonstrate the associated standard error. The markers are coloured differently by ethnicity.

Among the identified candidate SNPs, we observed a positive effect on cardiovascular traits for SNP rs10069648, along with 1,925 additional SNPs, in both the AFR and EUR populations. However, the significance of rs10069648 was notably higher in the AFR cohort (β = 5.5; MTAG p = 3.8 × 10^−8^) compared to the EUR population (β = 3.9; p = 8.3 × 10^−5^). Interestingly, rs10069648 was identified as a common variant in both the AFR (minor allele frequency [MAF] = 0.44) and EUR (MAF = 0.44) populations, suggesting its potential contribution to trait similarities between these populations. In addition, we identified 2,010 candidate SNPs that exhibited negative effect directions on cardiovascular traits in both the AFR and EUR populations.

Overall, our findings demonstrate the population-specific effects of SNPs located within the same genomic risk loci on cardiovascular traits. This highlights the differential influences at the SNP level while also indicating the presence of shared genetic loci across the AFR and EUR cohorts. Our findings emphasize the necessity of accounting for population variation in genetic studies to enhance precision in risk prediction and therapeutic strategies ^49,50^.

### The novelty of loci and variants associated with cardiovascular function

In a subsequent step, we aimed to identify MTAG known and novel SNPs and genomic risk loci associated with cardiovascular traits within AFR and European EUR populations, extending to SNPs in high LD within a 500kb LD block (refer to the Methods section for details). Simultaneously, we attempted to pinpoint novel SNPs linked to traits capable of modifying, affecting, or exhibiting associations with cardiovascular function, such as weight and body mass index.

Among the 50 significant genomic risk loci identified in the AFR population, we discovered 43 novel loci that have not been previously associated with these traits. Additionally, we observed 7 loci that were already known to be associated with the traits under study (refer to Supplementary Data 2 for comprehensive information on the novel and known loci). In contrast, out of the 829 loci identified in the EUR population, we found 47 novel loci that had not been previously linked to these traits. Notably, many loci (782) were already known to be associated with the traits (refer to Supplementary Data 2 for further details).

Focusing on the AFR cohort, among the 52 lead SNPs, our analysis revealed 45 novel variants (see methods for the definition of the novel). This group includes rs56195233 (Figure 4a), rs3019892 (Figure 4b), and rs74265878 (refer to Table 2 and Supplementary Data 3 for detailed information).

**Figure 4:**
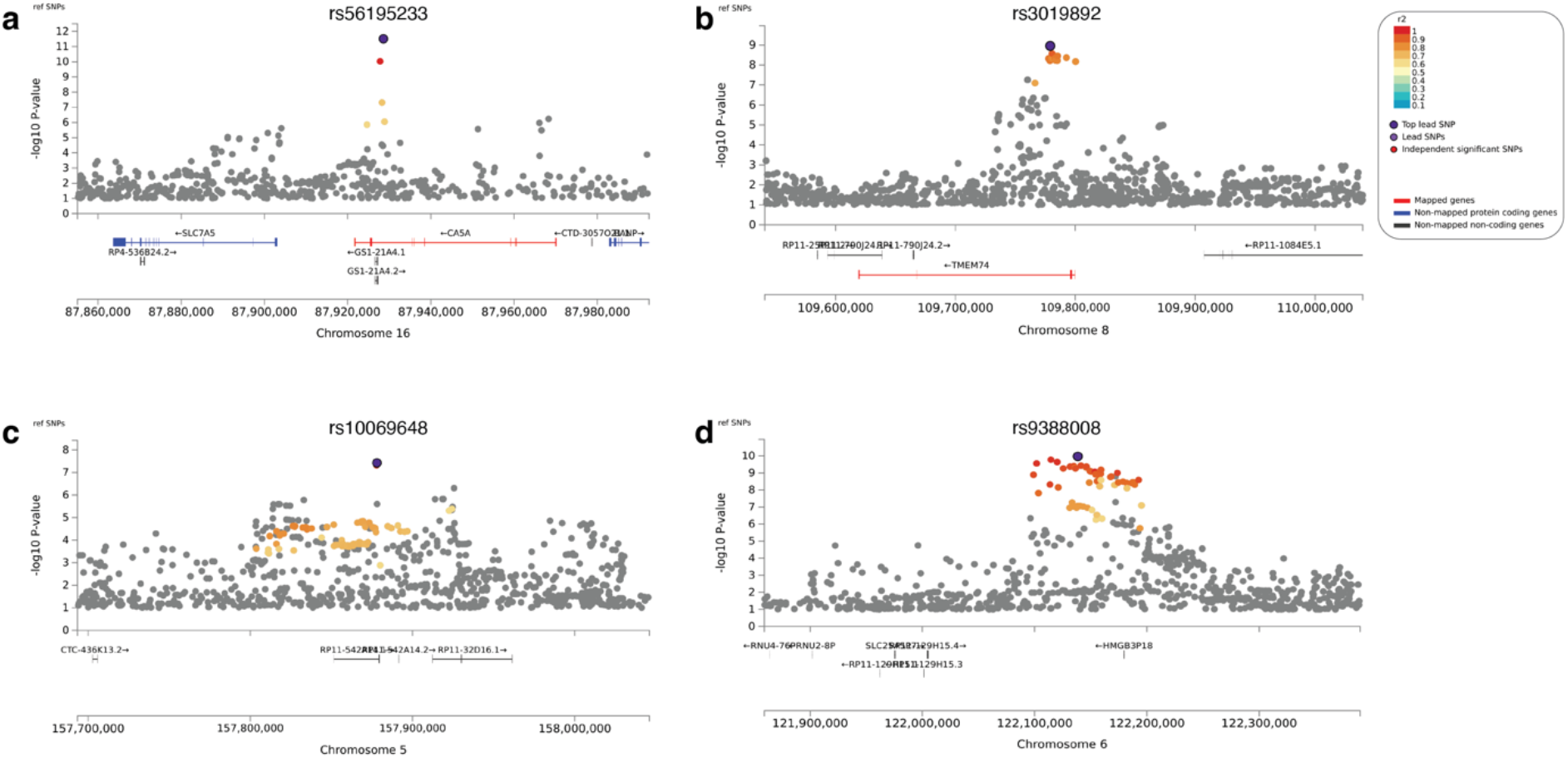
Regional Association Plots Highlighting Novel and Previously Identified MTAG Loci Associated with Cardiovascular Traits in African Population: The lead SNPs (a) rs56195233 and (b) rs3019892 represent novel findings, while the lead SNPs (c) rs10069648 and (d) rs9388008 have been previously linked to cardiovascular traits. Genes within the chromosomal loci are depicted in the lower panel. The filled circles indicate the position of the SNPs across the region on the x-axis and the negative logarithm of the association p-value on the y-axis. The lead SNP is highlighted in purple, with other SNPs within the locus coloured based on their linkage disequilibrium correlation value (r^2^) with the lead SNP, utilizing the African HapMap haplotype from the 1000 Genomes Project.

**Table 2:** Known and novel variants associated with cardiovascular traits.

| Population | Cardiovascular trait | Cardiovascular Associated Traits | Novel |
| --- | --- | --- | --- |
| Africans | 6 | 1 | 45 |
| Europeans | 724 | 185 | 947 |

In addition to these novel findings, we successfully identified 6 SNPs that have been previously associated with cardiovascular traits. These include rs10069648 (Figure 4c), rs9388008 (Figure 4d), and rs77199928. Specifically, rs10069648 has previously been linked with systolic, diastolic, and pulse pressure^51^, while rs9388008 has known associations with various traits such as heart rate, atrial fibrillation, and resting heart rate^51,52^.

Moreover, we detected one variant, rs116055168, associated with traits that modify or are interconnected with cardiovascular function, such as height and body mass index^17^.

We discovered 947 novel lead SNPs associated with cardiovascular traits for the EUR cohort. Out of these, 3:169171347:G:T (Figure 5a) and rs11330240 (Figure 5b) particularly stand out due to their strong associations (for comprehensive details, refer to Table 3 and Supplementary Data 3).

**Figure 5:**
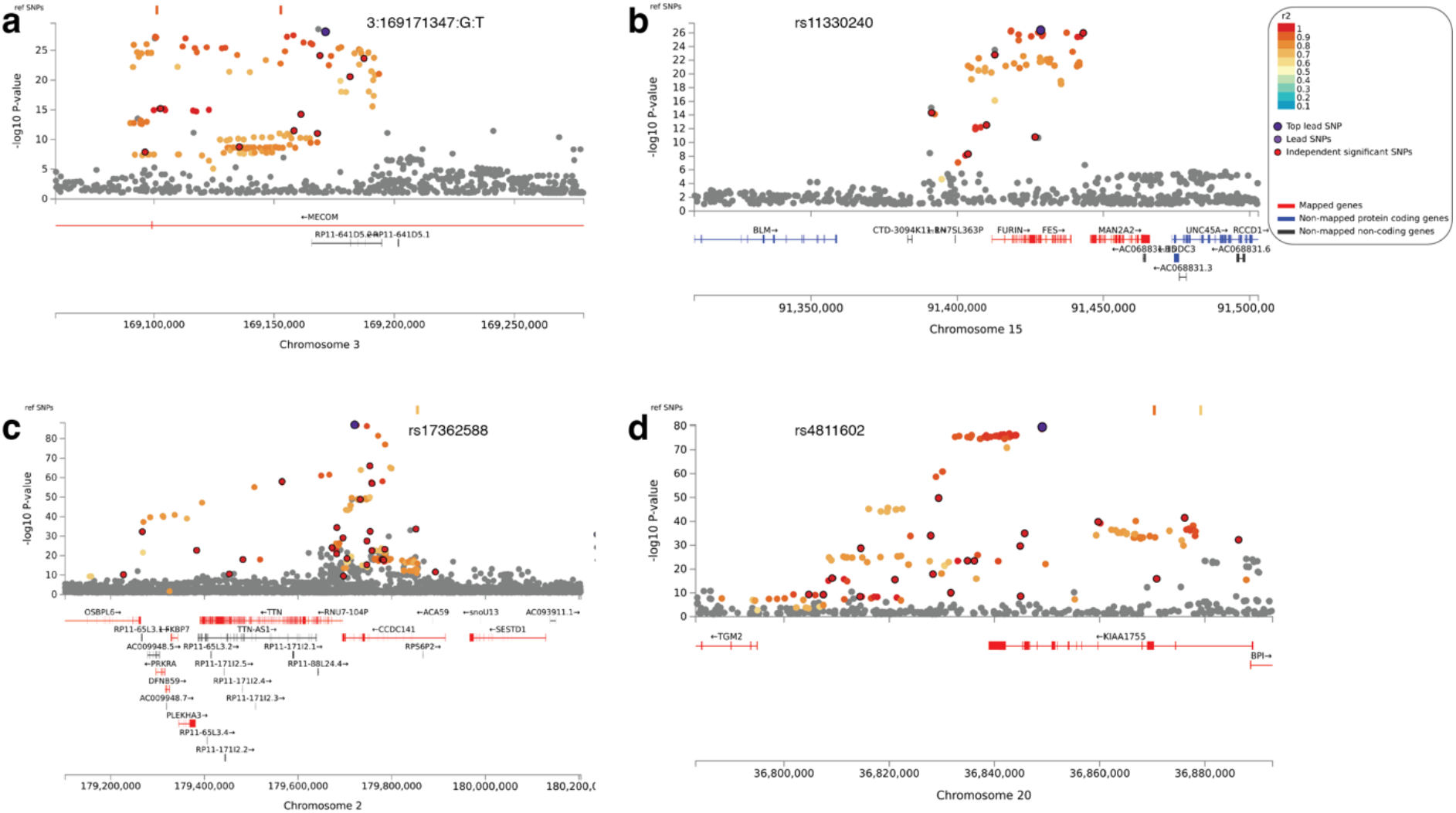
Regional Association Plots Highlighting Novel and Previously Identified MTAG Loci Associated with Cardiovascular Traits in European Population: Regional association plots for MTAG loci significantly associated with cardiovascular traits in the African population. The lead SNPs (a) 3:169171347:G:T and (b) rs11330240 represent novel findings, while the lead SNPs (c) rs17362588 and (d) rs4811602 have been previously linked to cardiovascular traits. Genes within the chromosomal loci are depicted in the lower panel. The filled circles indicate the position of the SNPs across the region on the x-axis and the negative logarithm of the association p-value on the y-axis. The lead SNP is highlighted in purple, with other SNPs within the locus coloured based on their linkage disequilibrium correlation value (r^2^) with the lead SNP, utilizing the European HapMap haplotype from the 1000 Genomes Project.

In addition to these novel discoveries, we confirmed the association of 724 previously identified SNPs related to cardiovascular traits. Among these, rs17362588 (Figure 5c), rs4811602 (Figure 5d), and rs10774625 have particularly strong associations. For instance, rs17362588 has been previously linked with diastolic blood pressure, pulse rate^51^ and hypertrophic cardiomyopathy^53^, while rs4811602^54,55^ and rs10774625^51^ have known associations with traits such as heart rate and blood pressure.

Furthermore, we highlighted 185 variants with known association with traits that modify or are interconnected with cardiovascular function. This includes rs11379773^56^ (Supplementary Figure 2e), rs5848907^57^ (Supplementary Figure 2f), and rs56057703^58^, which have been previously linked to body mass index and body weight, traits that indirectly impact cardiovascular function.

Our findings support the rich diversity of genetic variants that play critical roles in cardiovascular function, warranting further detailed investigation to unravel their full potential for risk prediction or informing treatment.

### Frequency of variants between Africans and Europeans

Our analysis of SNP frequency between AFR and EUR populations demonstrates notable differences in the distribution of genetic variants. Among the MTAG lead SNPs, we identified significant differences in SNP frequencies, confirming the potential influence of genetic background on cardiovascular traits (see Supplementary Figure 3).

Our study identified several SNPs with varying frequencies between the EUR and AFR populations. The SNPs with the highest frequency in the EUR population were rs2267666 (frequency = 0.76), rs2267667 (0.76), and rs7751481 (0.76). Conversely, the SNPs most frequent in the AFR population were rs4468 (0.81), rs10096421 (0.89), and rs35499486 (0.88). These differences highlight the distinct genetic backgrounds between the populations and underscore the importance of considering ancestry when analysing genetic data.

Additionally, our analysis uncovers numerous SNPs that show significant associations with cardiovascular traits exclusively within one population and not the other, highlighting the presence of population-specific genetic influences on cardiovascular health. These SNPs exhibit substantial frequency disparities between the EUR and AFR populations. For example, the observed frequency differences in these SNPs—such as rs2267666, rs2267667, and rs7751481 being more prevalent in the EUR population, and rs4468, rs10096421, and rs35499486 predominating in the AFR population—suggest that these genetic variants could have divergent effects on cardiovascular traits, potentially contributing to population-specific disease risk profiles and outcomes ^59,60^.

### Functional impact of the SNPs associated with the cardiovascular traits

We aimed to assess the functional impact of the SNPs associated with cardiovascular traits in AFR and EUR populations. By conducting functional mapping and annotation using FUMA, we uncovered unique patterns of SNP distribution in each population (Figure 6).

**Figure 6:**
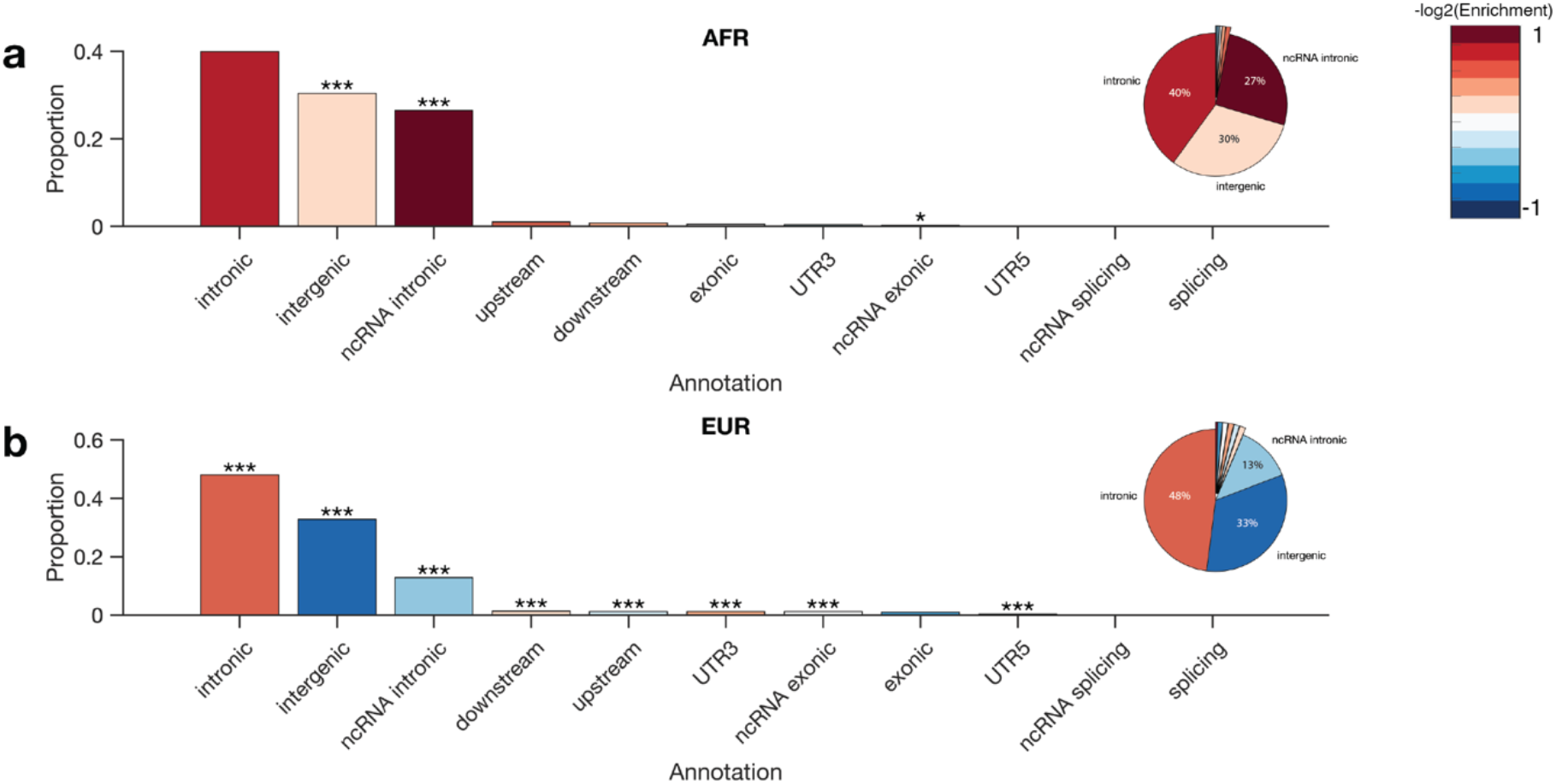
Functional Annotation of SNPs in AFR and EUR Populations. (a) African (AFR) population SNP annotations with corresponding proportions shown in the bar graph and pie chart. The bar graph indicates the proportion of SNPs in various genomic regions, with asterisks denoting significance levels: ***p<0.001, **p<0.01, *p<0.05. The pie chart shows the percentage distribution of SNPs across intronic, intergenic, and ncRNA-intronic regions. (b) European (EUR) population SNP annotations, presented in the same format as the AFR population. The significance levels are marked similarly on the bar graph, and the pie chart reflects the distribution within intronic, intergenic, and ncRNA-intronic regions. The heat map legend on the right side represents the -log2(Enrichment) value of SNPs in each annotation category, with 1 indicating the highest level of enrichment.

Across both populations, most of the SNPs were in non-coding regions, particularly in intergenic and intronic regions. However, there were notable differences in the distribution of SNPs within these regions between the AFR (Figure 6a) and EUR (Figure 6b) populations. In the EUR population, there was a higher proportion of SNPs in intronic regions (47.00% in EUR versus 39.68% in AFR), which was supported by a significantly higher enrichment in the EUR population (enrichment=1.29, p<0.0001).

Furthermore, the EUR population exhibited a higher proportion and significant enrichment of SNPs in untranslated regions (UTRs), specifically UTR3 (1.18% in EUR versus 0.64% in AFR) and UTR5 (0.40% in EUR versus 0.32% in AFR) (Figure 7). Additionally, there was a higher proportion of SNPs in downstream regions (1.34% in EUR versus 0.64% in AFR), non-coding RNA (ncRNA) exonic regions (1.16% in EUR versus 1.28% in AFR), and upstream regions (1.17% in EUR versus 0.72% in AFR) in the EUR population, suggesting the potential importance of these regions in cardiovascular traits within this group.

**Figure 7:**
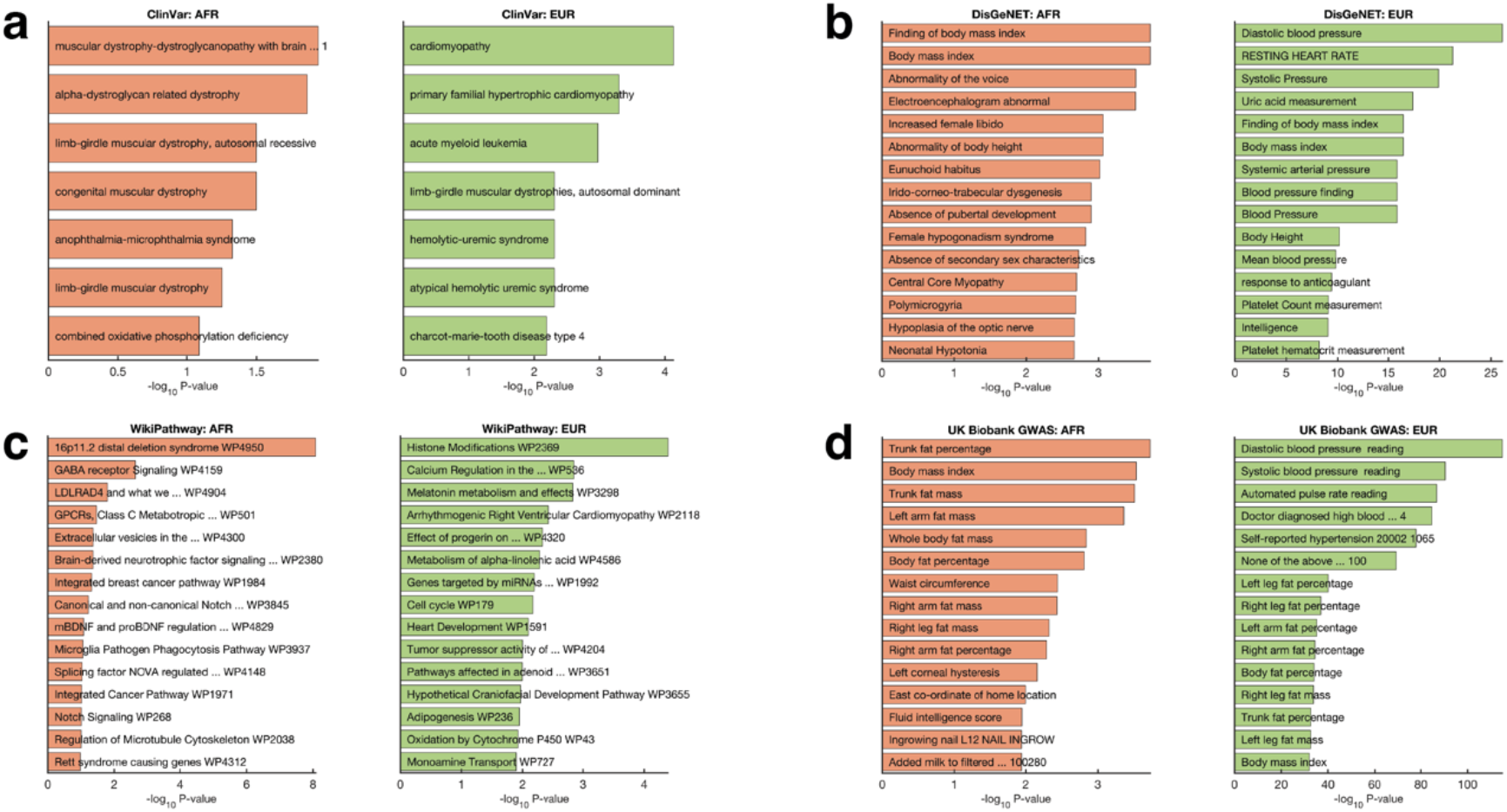
Functional and Pathway Enrichment Analyses: Plots illustrating enrichment analyses of (a) ClinVar diseases, (b) DisGeNET genes, (c) WikiPathway gene sets, and (d) UK Biobank GWAS terms. The analyses focus on genes mapped from SNPs using FUMA based on MTAG analysis results for individuals of African and European ancestry. Comprehensive *details of functional and* pathway enrichment results can be found in Supplementary Data 4.

Intergenic regions were significantly enriched in both populations, particularly in the EUR population (34.02% in EUR versus 46.52% in AFR), suggesting a potentially higher functional relevance of these regions in cardiovascular traits or reflecting population-specific genomic characteristics. Additionally, there was an absence of SNPs in splicing regions and a lower representation in exonic regions in both populations (1.03% in EUR versus 0.48% in AFR), highlighting the potentially deleterious effects of such variations.

Overall, our findings elucidate distinct patterns of SNP distribution in the AFR and EUR populations, providing valuable insights into the genetic architecture of cardiovascular traits across different ethnic groups.

### Differential genes and functional enrichments in Africans and Europeans

We employed FUMA^34^ to map the MTAG significant SNPs to distinct genes in each population. Here, we identified 1,267 candidate SNPs in the AFR group and a staggering 155,244 in the EUR group.

When analysing the mapped genes, we again found substantial population-specific genetic associations, with 3,011 genes uniquely associated within the EUR group and 36 distinct genes from the AFR group (Supplementary Figure 4). Only 10 genes were shared across both populations, suggesting their potential pan-ethnic influence on these traits (Supplementary Data 4).

Further, we utilised Enrichr^39^ to perform gene enrichment analysis within each group, identifying unique enriched pathways for the AFR and EUR populations. From the ClinVar database, the top three significant pathways in the AFR population were ‘muscular dystrophy-dystroglycanopathy’ (p=0.011), ‘alpha-dystroglycanopathy related dystrophy’, and ‘congenital muscular dystrophy’. In contrast, the EUR group showed significant enrichment in cardiomyopathy (p=7.4 ×10^−05^), ‘primary familial hypertrophic cardiomyopathy, and ‘acute myeloid leukaemia (Figure 7a).

When using DisGeNET^41^ for pathway analysis, we found the AFR population to be significantly associated with ‘body mass index’ (p=0.0002), ‘Finding of body mass index’, and ‘Abnormality of the voice’ pathways. For the EUR group, the top enriched pathways included diastolic blood pressure (p=7.9 × 10^−27^), resting heart rate, and systolic blood pressure (Figure 7b).

In the WikiPathway^42^ analysis, for the AFR group, the enrichment was highly related to ‘16p11.2 distal deletion syndrome’ (p= 8.4 × 10^−09^), ‘GABA receptor signalling’, and ‘LDLRAD4 and what we know about it’. Meanwhile, the EUR group demonstrated a strong association with ‘Histone modifications’ (4.1 ×10^−05^), ‘Calcium regulation in the cardiac cell’, and ‘Melatonin metabolism and effects’ (Figure 7c).

Finally, we performed enrichment analyses based on the UK Biobank gene as the background database^39^. In the AFR population, the top five significantly enriched terms were ‘Trunk fat percentage’ (p = 1.9 × 10^−5^), ‘Body mass index’ (p = 2.9 × 10^−5^), and ‘Trunk fat mass’ (p = 3.1 × 10^−5^). Conversely, the EUR population exhibited enrichment for cardiovascular traits, with the highest ranking being ‘Diastolic blood pressure’ (p = 1.3 x 10^-115^), ‘Systolic blood pressure’ (p = 3.5 × 10^−91^), and ‘Automated pulse rate reading’ (p = 1.7 × 10^−87^). Refer to Figure 7d and Supplementary Data 4 for more details.

These divergent pathway enrichments between the AFR and EUR populations suggest differences in the mechanisms through which genetic ancestry impacts trait manifestations, highlighting the necessity for a population-specific approach to understanding the genetic underpinnings of cardiovascular function. Additionally, shared genes among both populations suggest some potential pan-ethnic contributors to these traits, warranting further exploration and validation.

## Discussion

Our study presents a comprehensive analysis of the genetic variants associated with cardiovascular traits across AFR and EUR populations in the UKB. Our findings highlight the substantial genetic diversity underlying these traits in different ethnic groups, revealing both shared and unique genetic determinants that can contribute to disparities in cardiovascular health outcomes.

The identification of novel SNPs associated with cardiovascular traits in both the AFR and EUR populations substantiates the power of MTAG to reveal novel genetic associations. Previous studies have employed MTAG to elucidate the genetic architecture of complex traits^61^, and our work extends these findings by identifying 45 novel variants in the AFR population and an impressive 957 novel variants in the EUR population.

These discoveries highlight the genetic diversity underlying traits and emphasize the importance of more diverse and inclusive genomic research. Previous literature has noted that a disproportionate focus on European-ancestry populations in genetic studies has limited our understanding of disease genetics across diverse populations^18^. Importantly, our findings caution against the uncritical application of genetic risk prediction models across populations, emphasizing the need for population-specific genetic research in precision medicine^62^.

In addition, our study reveals remarkable differences in SNP frequencies between the AFR and EUR populations. These disparities, in line with previous findings^63–66^, highlight the complex interplay between genetic background and cardiovascular health. However, the discovery of shared genetic factors among AFR and EUR populations hints at some shared genetic aetiology for specific traits, a finding that could potentially be leveraged for broad therapeutic interventions^67^. Nevertheless, the significant number of unique genes identified in each population highlights the influence of population-specific genetic factors, likely due to varying population history, environmental interactions, or statistical factors, such as sample size^36,62,68,69^.

Functional mapping and annotation of the SNPs reveal unique patterns of SNP distribution in non-coding regions across the AFR and EUR populations. The increasing recognition of the importance of non-coding regions in the regulation of gene expression and disease susceptibility^70,71^ is supported by our findings. Notably, the associations spanned multiple traits, suggesting pleiotropic effects where a single gene or SNP can influence multiple traits^50^. An example of such a phenomenon is the SNP rs10069648, which we found linked to diverse traits, including blood pressure regulation, medication use, and longevity^51,57^. However, it is important to note that further studies are necessary to understand the biological mechanisms driving these associations^49^.

The enrichment of distinct functional pathways in each population highlights potential divergences in the pathophysiology of cardiovascular diseases, indicative of a complex interplay between genetic and environmental factors that differ across populations. For instance, in the EUR population, we identified pathways associated with cardiomyopathy and limb-girdle muscular dystrophy, both of which have previously been associated with cardiovascular traits^72,73^. Conversely, in the AFR population, BMI, body fat, enrichment of rasopathy and oxidative phosphorylation deficiency pathways were observed, which might suggest unique molecular mechanisms underlying cardiovascular disease in this population^74,75^. The significant enrichment of the identified variants within gene sets known to influence cardiovascular parameters, such as blood pressure, pulse rate, and heart rate, in the EUR population provides further evidence of a nuanced genetic landscape. Notably, this enrichment aligns well with previously conducted GWAS, thus supporting our findings. However, this also highlights the extensive research focus on the EUR population in cardiovascular studies, emphasizing the need for greater efforts in investigating underrepresented populations, such as the AFR group.

Overall, our results, in which we found different variants associated with cardiovascular traits at the common genomic risk loci in AFR and EUR populations, are supported by the fact that a SNP can have different effects on a trait in different populations due to various factors. For example, a SNP that is associated with a trait in one population may not be associated with the same trait in another population if the SNP has a low frequency or is absent^76^, is in LD with a causal variant in one population but not the other^77^, interacts with different environmental factors^78^, or has a different genetic background or epistatic effects with other SNPs that modulate the trait^79^.

Our study is not without limitations. The functional implications and mechanistic roles of the identified genetic variants need further validation. This caveat is common to many GWAS studies and underlines the importance of combining GWAS with functional studies for a more comprehensive understanding of the genotype-phenotype relationship^49^. Moreover, the smaller sample size of the AFR population may have limited our ability to identify all associated variants, echoing calls for larger and more diverse genetic studies^80^.

In conclusion, our findings add to the growing body of literature describing the complexity and diversity of the genetic underpinnings of cardiovascular traits across different populations^81–83^. This work lays a foundation for more precise and personalized approaches to preventing and managing cardiovascular diseases, ultimately aiming to reduce health disparities across different ethnic groups.

## Data Availability

The datasets that support the results presented in this manuscript are available from: the UK Biobank; https://www.ukbiobank.ac.uk, dbSNP; https://www.ncbi.nlm.nih.gov/snp, and the GWAS Catalog; https://www.ebi.ac.uk/gwas.

## Code Availability

The code to reproduce most of the results presented here is available from the corresponding authors upon request.

## Acknowledgements

The funding for this project was provided by H3ABioNet, supported by the National Institutes of Health Common Fund under grant number U24HG006941. The content of this publication is solely the authors’ responsibility and does not necessarily represent the official views of the National Institutes of Health.

## Author Contributions

M.S., S.E., and N.M. conceptualized the study. M.S., N.M., S.E., and J.C. designed the methodology, and M.M. M.S., M.M., J.C., and S.E. formally analysed the data. M.S., N.M., and S.E. drafted the manuscript. Editing and reviewing the manuscript were carried out by M.S., N.M., S.E., J.C., and M.M. Data visualisations were produced by M.S. and S.E.

## Description of Additional Supplementary Files

**Supplementary Data 1:** Supplementary data of genome-wide associations of cardiovascular traits. The spreadsheet contains the following results/datasets according to the sheet name. “Welch Test Results” presents all the test statistics for the Welch test comparing cardiovascular measures (SBP, DBP, Pulse Rate and MHR) among African and European populations. “Variable Stats” present the descriptive statistics of the cardiovascular trait measures.

**Supplementary Data 2:** Cardiovascular trait MTAG significant variants and SNPs in African and European populations: This file has multiple sheets that further illustrate and explain the study results. The “EUR - Lead SNPs” and “AFR - Lead SNPs” sheets present the top SNPs associated with cardiovascular traits in both populations, further highlighting the differential and shared genetic influences on these traits. The “EUR - Genomic Risk Loci” and “AFR - Genomic Risk Loci” sheets contain a comprehensive list of genomic risk loci identified in the European and African cohorts. Finally, the “Beta Comparison” sheet provides detailed comparisons of beta estimates for all investigated SNPs in both populations, offering a granular view of their effects.

**Supplementary File 3:** The spreadsheet provides more specific details about the identified SNPs and their association with cardiovascular traits in the African and European cohorts. The “AFR Known Catalog Traits” sheet includes information extracted from the GWAS Catalog regarding known cardiovascular trait variants found among the SNPs in the African cohort. The “AFR Lead SNP Annon” sheet provides an annotation of the lead SNPs in the African cohort, distinguishing between novel variants, those already associated with cardiovascular traits, and those known to be variants for these traits. Similarly, the “EUR Known Catalog Traits” sheet gives an annotation of the lead SNPs in the European cohort, differentiating novel variants, those previously associated with cardiovascular traits, and those known to be variants for these traits. Finally, the “AFR Lead SNP Annon” sheet presents information from the GWAS Catalog about known cardiovascular trait variants among the SNPs in the European cohort.

**Supplementary File 4:** Integrated enrichment analysis results present a comprehensive functional enrichment analysis of the genes within which the identified MTAG significant SNPs are mapped and organised according to the African and European cohorts. Each sheet in this file is dedicated to the enrichment results derived from a specific database and population cohort, providing a targeted exploration of the functional implications of the SNPs. These sheets are titled according to the respective database and population, such as “ClinVar - AFR” and “ClinVar - EUR”, and similarly for ‘DisGeNET’, ‘UK Biobank GWAS’, and ‘WikiPathway’.

