## Supplementary Information for "Multitrait Genome-Wide Analysis in the UK Biobank Reveals Novel and Distinct Variants Influencing Cardiovascular Traits in Africans and Europeans"

***Supplementary Table 1:*** *Single-trait GWAS statistics of SNPs associated with cardiovascular traits in AFR and EUR populations.*

| Trait | Ancestry | Genomic Risk Loci | Lead SNPs | Ind. Sig. SNPs | Candidate SNPs | Candidate GWAS Tagged SNPs | Mapped Genes | |
| --- | --- | --- | --- | --- | --- | --- | --- | --- |
| Pulse Rate | AFR | 2 | 2 | 5 | 80 | 80 | | 0 |
| Diastolic BP | AFR | 0 | 0 | 0 | 0 | 0 | | 0 |
| Systolic BP | AFR | 0 | 0 | 0 | 0 | 0 | | 0 |
| Max Heart Rate | AFR | 0 | 0 | 0 | 0 | 0 | | 0 |
| Diastolic BP | EUR | 642 | 1090 | 2988 | 101698 | 101373 | | 2281 |
| Max Heart Rate | EUR | 25 | 26 | 36 | 913 | 913 | | 41 |
| Pulse Rate | EUR | 583 | 1224 | 3365 | 88845 | 88292 | | 2116 |
| Systolic BP | EUR | 590 | 980 | 2763 | 91192 | 90648 | | 2072 |

***Supplementary Table 2:*** *Multi-trait GWAS statistics of SNPs associated with cardiovascular traits in AFR and EUR populations.*

| Results Parameter | AFR | EUR |
| --- | --- | --- |
| Genomic Risk Loci | 50 | 829 |
| Lead SNPs | 52 | 1,856 |
| Individually Sig. SNPs | 54 | 5,416 |
| Candidate SNPs | 772 | 138,032 |
| Candidate GWAS Tagged SNPs | 753 | 134,477 |
| Mapped Genes | 46 | 3033 |

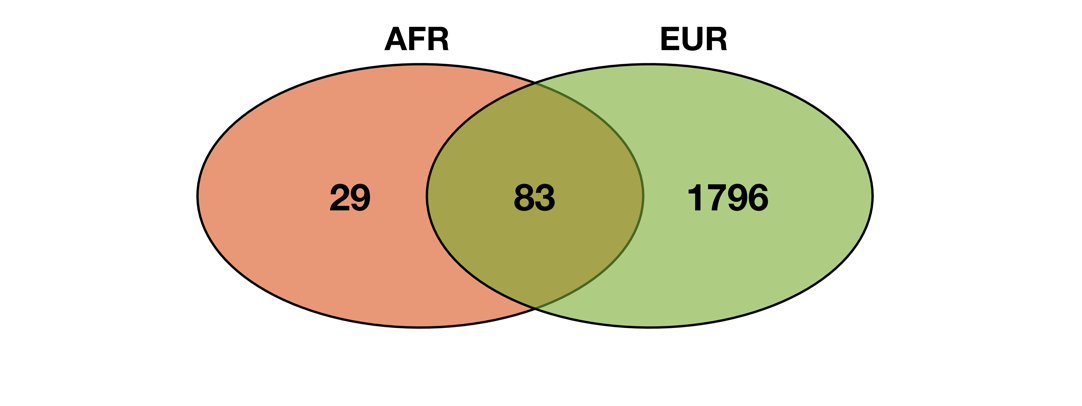

**Supplementary Figure 1:** Venn diagrams illustrating the overlap of lead variants associated with cardiovascular traits in African and European populations, as identified by our multi-trait GWAS analysis. The intersection comprises variants that are directly and significantly associated with cardiovascular traits in both populations, as well as those that are replicated from one population to the other.

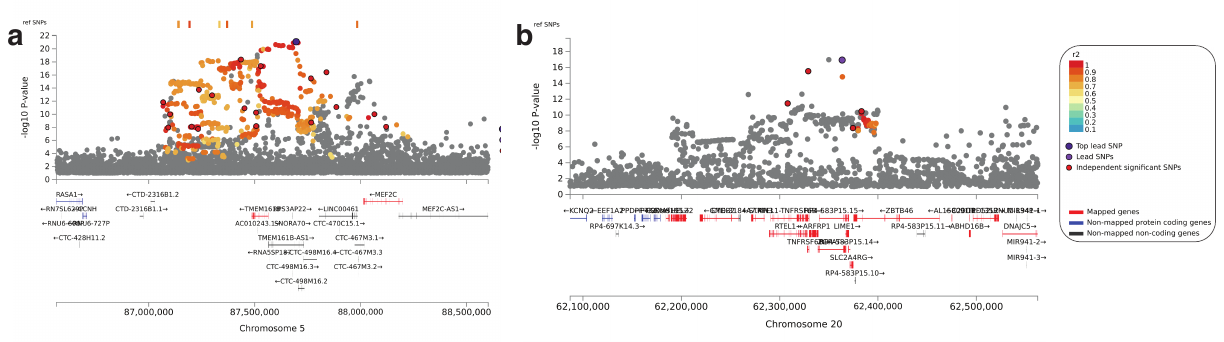

**Supplementary Figure 3:** Regional association plots for MTAG loci for the lead SNPs **(a)** rs11379773 and **(b)** rs5848907. The genes within the chromosomal loci are shown in the lower panel. The filled circles show the position of the SNPs along the region on the x-axis and the negative logarithm of the association p-value on the y-axis. The lead SNP is shown in purple, and the SNPs within the locus are coloured based on the linkage disequilibrium correlation value (r^2^) with the lead SNP based on the European HapMap haplotype from the 1000 genome project.

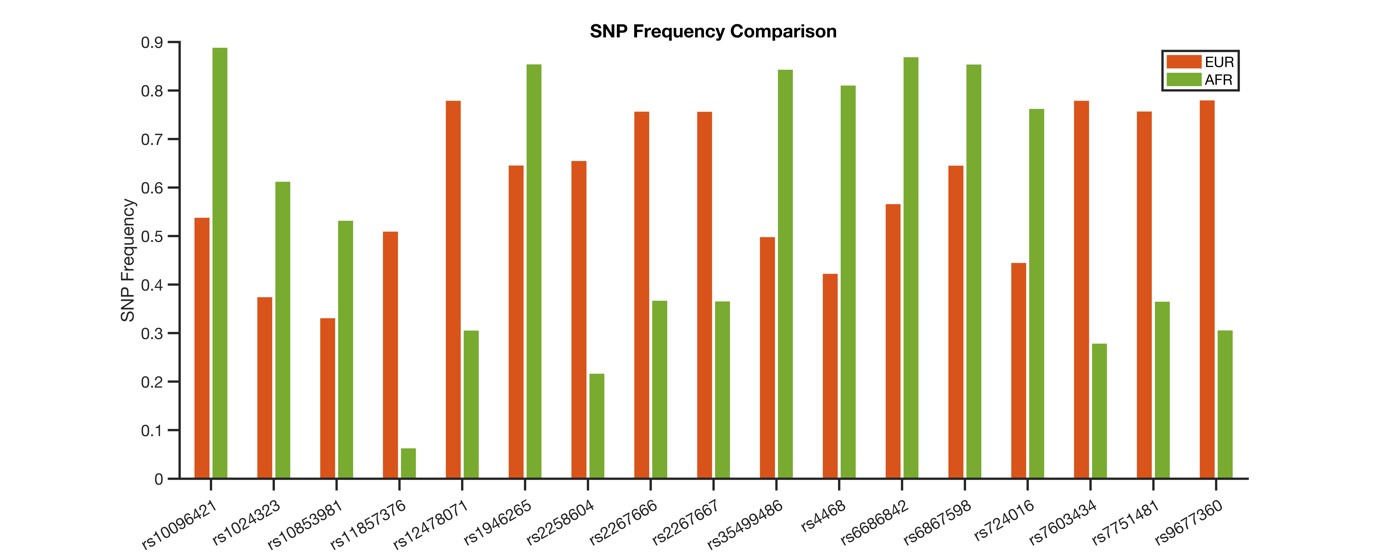

**Supplementary Figure 4:** Bar graph depicting SNPs linked to cardiovascular traits, which demonstrate the most pronounced differences in frequencies between Africans and Europeans. Notably, these SNPs may not be the most significantly associated with cardiovascular traits in the MTAG analysis among individuals of African and European ancestry.

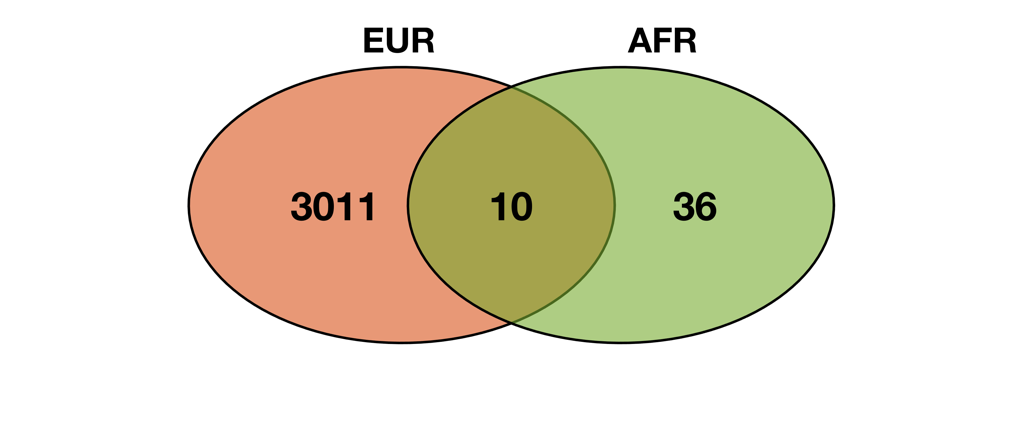

**Supplementary Figure 5:** Venn diagrams demonstrating the overlap of genes, identified via FUMA, in African and European populations from SNPs associated with cardiovascular traits, as determined through our multi-trait GWAS analysis.
